# Cross-sectional analysis of the humoral response after SARS-CoV-2 vaccination in Sardinian Multiple Sclerosis patients, a follow-up study

**DOI:** 10.1101/2022.05.19.22275317

**Authors:** Maria Laura Idda, Maristella Pitzalis, Valeria Lodde, Annalisa Loizedda, Jessica Frau, Monia Lobina, Magdalena Zoledziewska, Francesca Virdis, Giuseppe Delogu, Maria Giuseppina Marini, Maura Mingoia, Marco Masala, Lorena Lorefice, Marzia Fronza, Daniele Carmagnini, Elisa Carta, Silvy Pilotto, Paolo Castiglia, Paola Chessa, Sergio Uzzau, Gabriele Farina, Paolo Solla, Maristella Steri, Marcella Devoto, Edoardo Fiorillo, Matteo Floris, Roberto Ignazio Zarbo, Eleonora Cocco, Francesco Cucca

**Author notes:** **Correspondence:** Maristella Pitzalis, Maria Laura Idda. These authors have contributed equally to this work and share first authorship (Maria Laura Idda, Maristella Pitzalis). These authors have contributed equally to this work and share last authorship (Matteo Floris, Roberto Ignazio Zarbo, Eleonora Cocco, Francesco Cucca).

## Abstract

Monitoring immune responses to SARS-CoV-2 vaccination and its clinical efficacy over time in Multiple Sclerosis (MS) patients treated with disease-modifying therapies (DMTs) help to establish the optimal strategies to ensure adequate COVID-19 protection without compromising disease control offered by DMTs. Following our previous observations of the humoral response one month after two doses of BNT162b2 vaccine (T1) in MS patients differently treated, here we present a cross-sectional and longitudinal follow-up analysis six months following vaccination (T2, n=662) and a month following the first booster (T3, n=185). Consistent with results at T1, humoral responses were decreased in MS patients treated with fingolimod and anti-CD20 therapies compared with untreated patients also at the time points considered here (T2 and T3). Interestingly, a strong upregulation one month after the booster was observed in patients under every DMTs analyzed, including those treated with fingolimod and anti-CD20 therapies. And although patients taking these latter therapies had a higher rate of COVID-19 infection five months after the first booster, only mild symptoms that did not require hospitalization were reported for all the DMTs analyzed here. Based on these findings we anticipate that additional vaccine booster shots will likely further improve immune responses and COVID-19 protection in MS patients treated with any DMT.

## Introduction

General population data on SARS-CoV-2 vaccination support its effectiveness in preventing COVID-19 infection (1). Still, the magnitude of protection it offers to Multiple Sclerosis (MS) patients receiving certain disease-modifying therapies (DMTs) is not completely clear.

Several evidence have already demonstrated that the humoral and cellular response after SARS-CoV-2 vaccination were strongly affected by the treatment with certain DMTs used to ameliorate MS symptoms (2–4). Notably, azathioprine (AZA), fingolimod (FTY) and anti-CD20 treatments, including ocrelizumab (OCR) and rituximab (RTX), negatively influences the humoral response after SARS-CoV-2 vaccination and likely affect the level of protection against COVID-19 (2–4). Additionally, older age, male sex and active smoking were significantly associated with lower antibody titers against SARS-CoV-2 vaccine in MS patients (2,5).

Based on these results, it has been suggested that the immune response after SARS-CoV-2 vaccination can be improved through additional booster shots, and by optimally adjusting vaccination timing based on the timing of immune cell repopulation after the last administration of specific immunosuppressive DMTs (6). Specifically, in contrast to the initial international recommendation for timing DMTs (1) of a 3-month waiting time after the last dose of immune-suppressive therapies, we and others have recommended a 6-month waiting time (2,7). It is now important to further monitor, over time and after booster doses, the immune responses and clinical efficacy of SARS-CoV-2 vaccination in MS patients. This will provide new insight to define the most effective strategies that will ensure optimal treatment of MS providing at the same time the most effective prevention of COVID-19 and especially of its severe forms.

Following our initial observations of humoral response one month after two doses of BNT162b2 vaccine in MS patients treated with different DMTs or untreated (2) here we present a cross-sectional and longitudinal follow-up analysis on the humoral responses to BNT162b2 vaccination 6 months after the second dose, and a month after the third dose (booster). The effect of previous or concomitant detectable SARS-CoV-2 infection, as well as age, sex, and active smoking was also considered. Reciprocally, we provide preliminary evidence of the impact of vaccination on the incidence of SARS-CoV-2 infection and its clinical severity 5 months after the third dose of vaccine. The findings further help in defining an appropriate immunization strategy in MS patients in relation to DMTs and other factors influencing humoral immunity.

## Methods

### Study participants

847 MS patients who had received two BNT162b2 vaccine injections, 21 days apart, followed by a booster 6 month later were enrolled between October 2021 and January 2022 from the MS clinical centers of Cagliari and Sassari, Sardinia (Italy). Each injection contained 30μg of BNT162b2 (0.3ml volume per dose). 662 MS patients were analyzed 6 months after the second dose (T2) and 185 MS patients a month after the booster (third) dose (T3). Data from 912 MS patients at 4 weeks after the second dose (T1) were also used (2) (**Figure 1**).

**Figure 1.**
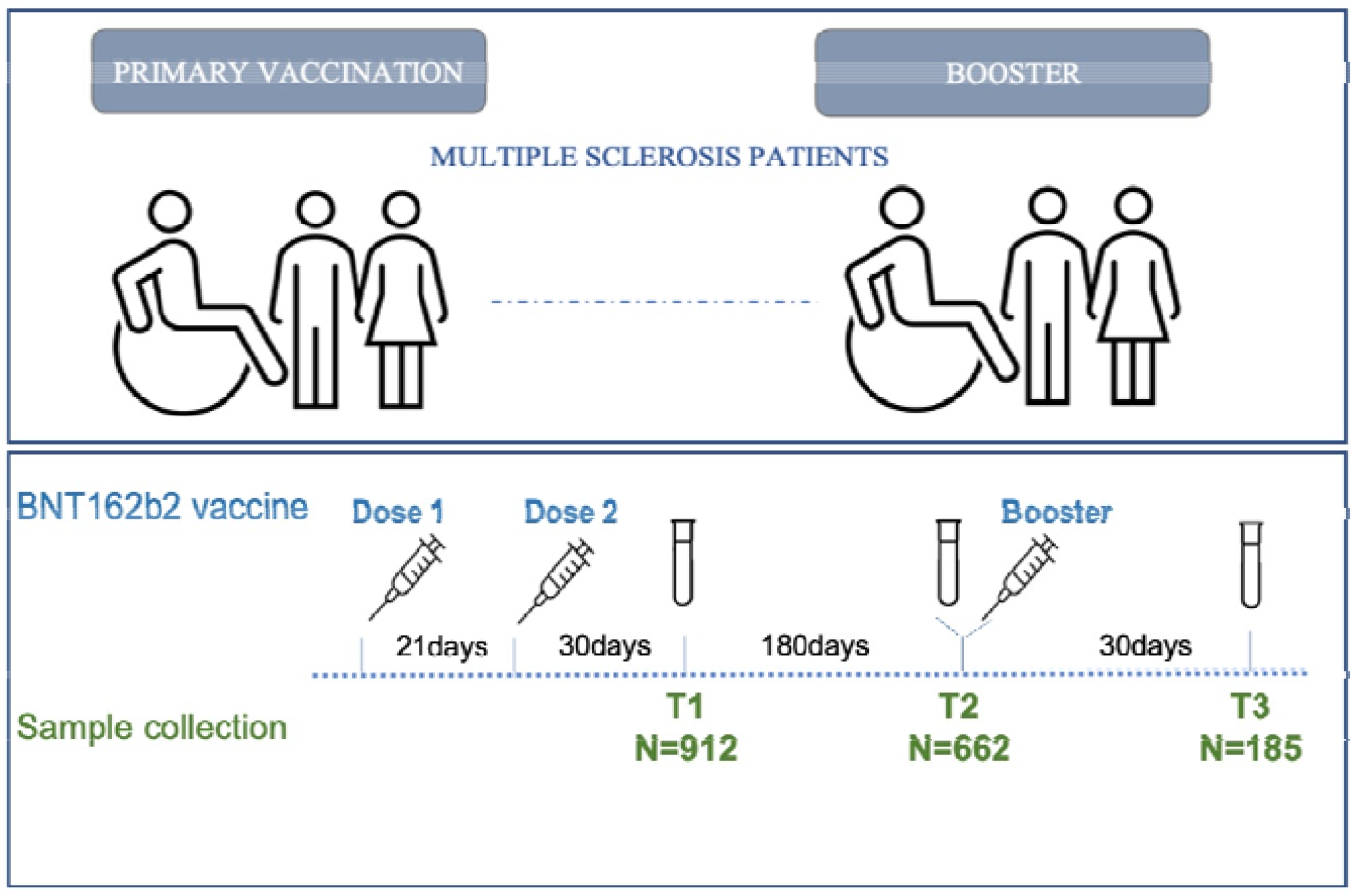
Timeline of MS patients’ enrollment. Schematic of the vaccination (blue) and sample collection (green) timeline in MS patients.

In summary we analyzed 1,944 serum samples of 1,307 unique MS patients. Of these, only 94 were recruited at each time point (T1, T2 and T3). The overlap between samples was 31.8% between T2 and T1, 22.8% between T2 and T3, and 13.9% between T3 and T1.

All MS patients, diagnosed according to the McDonald criteria, were contacted using different communication methods. They were questioned concerning previous SARS-CoV-2 infection and the manifestation of any adverse events following vaccination. Clinical and demographic information were collected for each patient, including age, sex, disability score, disease subcategory and disease-modifying treatment. All the data collected have been summarized in **Table 1**.

**Table 1.**
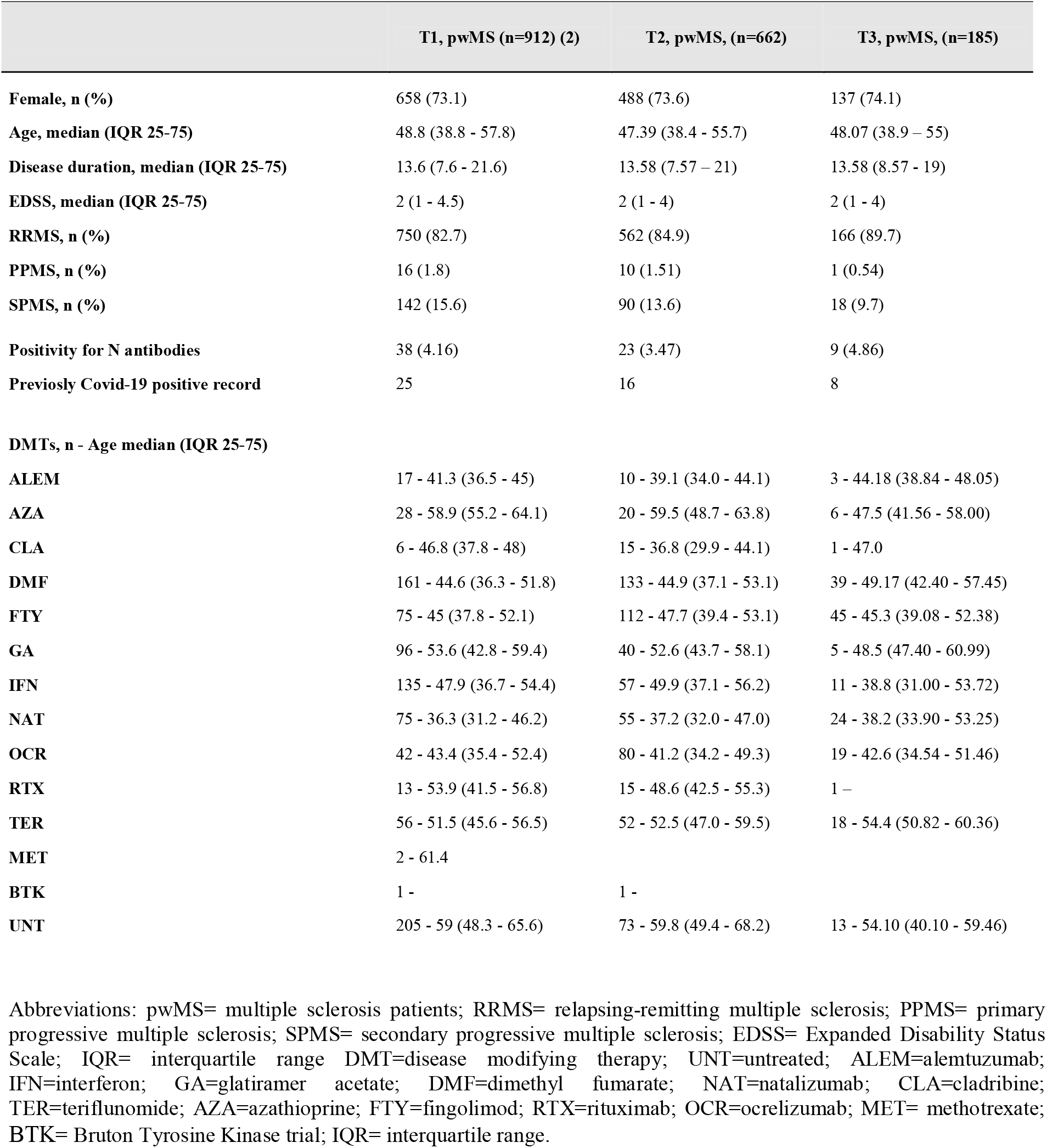
Clinical and demographic characteristics of multiple sclerosis patients at T1, T2 and T3.

### Detection of SARS-CoV-2 IgG antibodies

Blood samples were collected in vacutainer tubes containing clot activator and gel separator.

Samples were processed within two hours after blood collection to avoid time-dependent artifacts, and subsequently serum was stored at -80°C until use. Quantification of SARS-CoV-2 antibodies direct against the proteins Spike (S) or Nucleocapsid (N) in human serum was performed using the electrochemiluminescence immunoassays Elecsys® Anti-SARS-CoV-S and Elecsys® Anti-SARS-CoV-N (Roche). Anti-S and Anti-N results are expressed as units per ml (U/ml). The anti-N reactive index cutoff (COI) used is > 1.

### Statistical analysis

For all categorical variables the absolute number and percentage were reported, and for quantitative variables the median and interquartile range (IQR) were reported. Differences between patient groups - stratified according to therapy and selected for Anti-N antibody negativity - were assessed with a negative binomial generalized linear mixed-effects model, which takes into account the nature of the outcome variable (anti-S, non-negative count data); in addition to therapy, the models also consider the contribution of other variables such as age, smoke, sex, Expanded Disability Status Scale (EDSS), disease duration, and clinical sampling center. Results are presented as Incidence Rate Ratio, calculated as the exponential of the regression coefficient (8). Differences between medians were tested with the Mann-Whitney test. Differences between groups in longitudinal data were tested using the non-parametric Friedman test for repeated measures, followed by a post-hoc pairwise comparison using paired Wilcoxon signed-rank test. All statistical analyses were performed using R v.4.0.3 software with the following CRAN libraries: ggplot2, dplyr, readxl, MASS, kableExtra, rstatix. P values <0.05 were considered statistically significant.

### Ethics and data collection

The study was reviewed and approved by the Ethical Review Boards ATS Sardegna - Prot. N° 2492/CE. Patient data and samples were coded anonymously to ensure confidentiality during sample processing and data analysis. The patients/participants provided their written informed consent to participate in this study.

## Results

### Description of the studied MS cohort

A total of 1,944 sera were analyzed at three different time points following BNT162b2 vaccination. Results from 912 MS patients obtained 4 weeks after the second dose (T1) of vaccine have already been published (2) and have been used here as baseline for the two subsequent time points. 662 MS patients were analyzed 6 months after the second dose (T2) and 185 MS patients 4 weeks after the booster dose (T3) (**Figure 1**).

The T1 cohort has been previously described (2). The T2 MS cohort included 448 (73.6%) female and 214 (26.4%) male patients of whom 84.9% had relapsing-remitting MS (RR), 1.51% had primary progressive MS (PP), and 13.6% had secondary progressive MS (SP). Finally, the T3 MS cohort included 137 (74.1%) female and 48 (25.9%) male patients of whom 89.7% had RR, 0.54% had PP and 9.7% had SP (**Table 1**).

Untreated MS patients at T2 and at T3 were respectively 73 (11%) and 13 (7%). The remaining 589 (88.9%) at T2 and 172 (92.9%) at T3 were treated with different DMTs. The most common treatments were dimethyl fumarate (DMF) at T2 and FTY at T3. DMTs used less frequently included cladribine (CLA) and RTX, only one patient included in a study with a Bruton’s tyrosine kinase (BTK) inhibitor. No patients treated with methotrexate (MET) were present either at T2 or T3. A detailed description of the specific DMTs received by the MS patients along with their demographic characteristics at T1, T2 and T3 are summarized in **Table 1**.

### Disease-Modifying Therapies Impact Humoral Responses to BNT162b2 Vaccine

To evaluate the effects of different DMTs on humoral responses to BNT162b2 vaccine, we applied a negative binomial generalized linear mixed-effects model in patients negative for anti-N antibodies production for both T2 and T3 time points separately. Only treatments with data available for at least 10 patients were considered in this analysis.

In line with previously reported humoral responses to vaccine one month after the second dose (T1) (2,7), after six months (T2) we observed a significant difference in anti-S antibody levels between patients untreated (UNT) and those treated with FTY (IRR = 0.17, p = 1.82×10^−21^), OCR (IRR = 0.20, p = 4.54×10^−42^) and RTX (IRR = 0.33, p = 1.35×10^−17^). No significant difference was observed for the other DMTs (**Table 2 and Figure 2A**). Similarly, one month after the booster (T3) we observed a significantly lower level of anti-S antibodies in MS patients treated with FTY (IRR = 0.37, p = 1.52×10^−11^) and OCR (IRR = 0.34, p = 1.20×10^−14^) compared to untreated patients. Other treatments did not show significant results (**Table 3 and Figure 2B**).

**Table 2.**
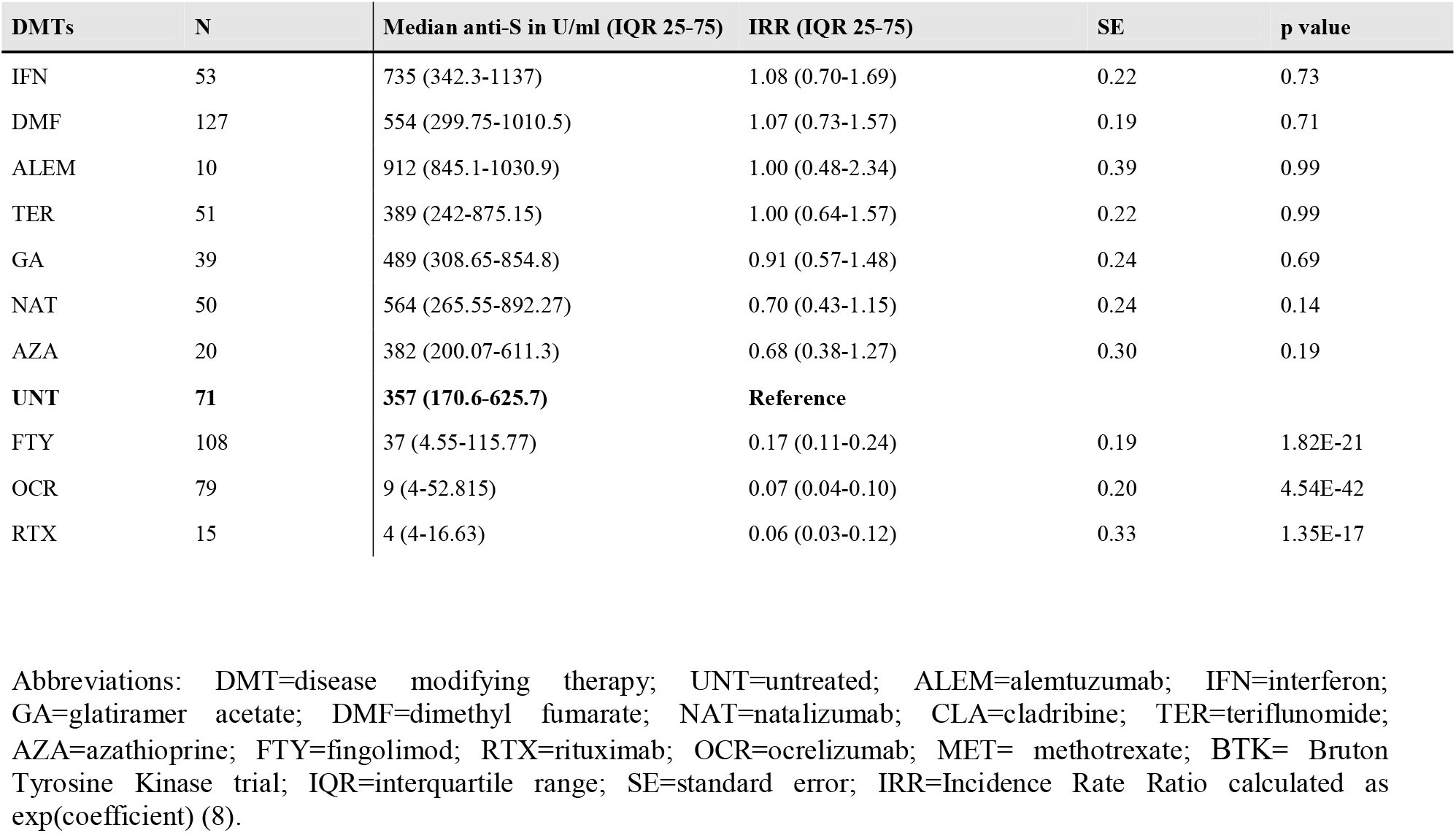
Negative binomial generalized linear mixed-effects model of anti-S-Ab levels in untreated and treated MS patients (anti-N negative) 6 months after the second dose (T2) of BNT162b2 vaccine.

| DMTs | N | Median anti-S in U/ml (IQR 25-75) | IRR (IQR 25-75) | SE | p value |
| --- | --- | --- | --- | --- | --- |
| IFN | 53 | 735 (342.3-1137) | 1.08 (0.70-1.69) | 0.22 | 0.73 |
| DMF | 127 | 554 (299.75-1010.5) | 1.07 (0.73-1.57) | 0.19 | 0.71 |
| ALEM | 10 | 912 (845.1-1030.9) | 1.00 (0.48-2.34) | 0.39 | 0.99 |
| TER | 51 | 389 (242-875.15) | 1.00 (0.64-1.57) | 0.22 | 0.99 |
| GA | 39 | 489 (308.65-854.8) | 0.91 (0.57-1.48) | 0.24 | 0.69 |
| NAT | 50 | 564 (265.55-892.27) | 0.70 (0.43-1.15) | 0.24 | 0.14 |
| AZA | 20 | 382 (200.07-611.3) | 0.68 (0.38-1.27) | 0.30 | 0.19 |
| **UNT** | **71** | **357 (170.6-625.7)** | **Reference** |  |  |
| FTY | 108 | 37 (4.55-115.77) | 0.17 (0.11-0.24) | 0.19 | 1.82E-21 |
| OCR | 79 | 9 (4-52.815) | 0.07 (0.04-0.10) | 0.20 | 4.54E-42 |
| RTX | 15 | 4 (4-16.63) | 0.06 (0.03-0.12) | 0.33 | 1.35E-17 |

Abbreviations: DMT=disease modifying therapy; UNT=untreated; ALEM=alemtuzumab; IFN=interferon; GA=glatiramer acetate; DMF=dimethyl fumarate; NAT=natalizumab; CLA=cladribine; TER=teriflunomide; AZA=azathioprine; FTY=fingolimod; RTX=rituximab; OCR=ocrelizumab; MET= methotrexate; BTK= Bruton Tyrosine Kinase trial; IQR=interquartile range; SE=standard error; IRR=Incidence Rate Ratio calculated as exp(coefficient) (8).

**Table 3.** Negative binomial generalized linear mixed-effects model of anti-S-Ab levels in untreated and treated MS patients (anti-N negative) one month after the BNT162b2 booster (T3).

| DMTs | N | Median anti-S in U/ml (IQR 25-75) | IRR (IQR 25-75) | SE | p value |
| --- | --- | --- | --- | --- | --- |
| IFN | 11 | 24639 (18339-89010) | 1.95 (0.75-5.08) | 0.48 | 0.16 |
| TER | 18 | 14008 (7791-24802) | 0.90 (0.36-2.15) | 0.43 | 0.80 |
| NAT | 21 | 16540 (9608-25000) | 0.76 (0.30-1.82) | 0.43 | 0.52 |
| DMF | 37 | 13346 (9031-25000) | 0.74 (0.32-1.60) | 0.39 | 0.44 |
| **UNT** | **12** | **16989 (12475-37407)** | **Reference** |  |  |
| FTY | 42 | 764 (315-2944) | 0.08 (0.04-0.17) | 0.37 | 1.52e-11 |
| OCR | 19 | 324 (8-1668) | 0.04 (0.01-0.09) | 0.43 | 1.20e-14 |

Abbreviations: DMT=disease modifying therapy; UNT=untreated; IFN=interferon; DMF=dimethyl fumarate; NAT=natalizumab; TER=teriflunomide; FTY=fingolimod; RTX=rituximab; OCR=ocrelizumab; IQR= interquartile range; SE= standard error; IRR=Incidence Rate Ratio calculated as exp(coefficient) (8).

**Figure 2.**
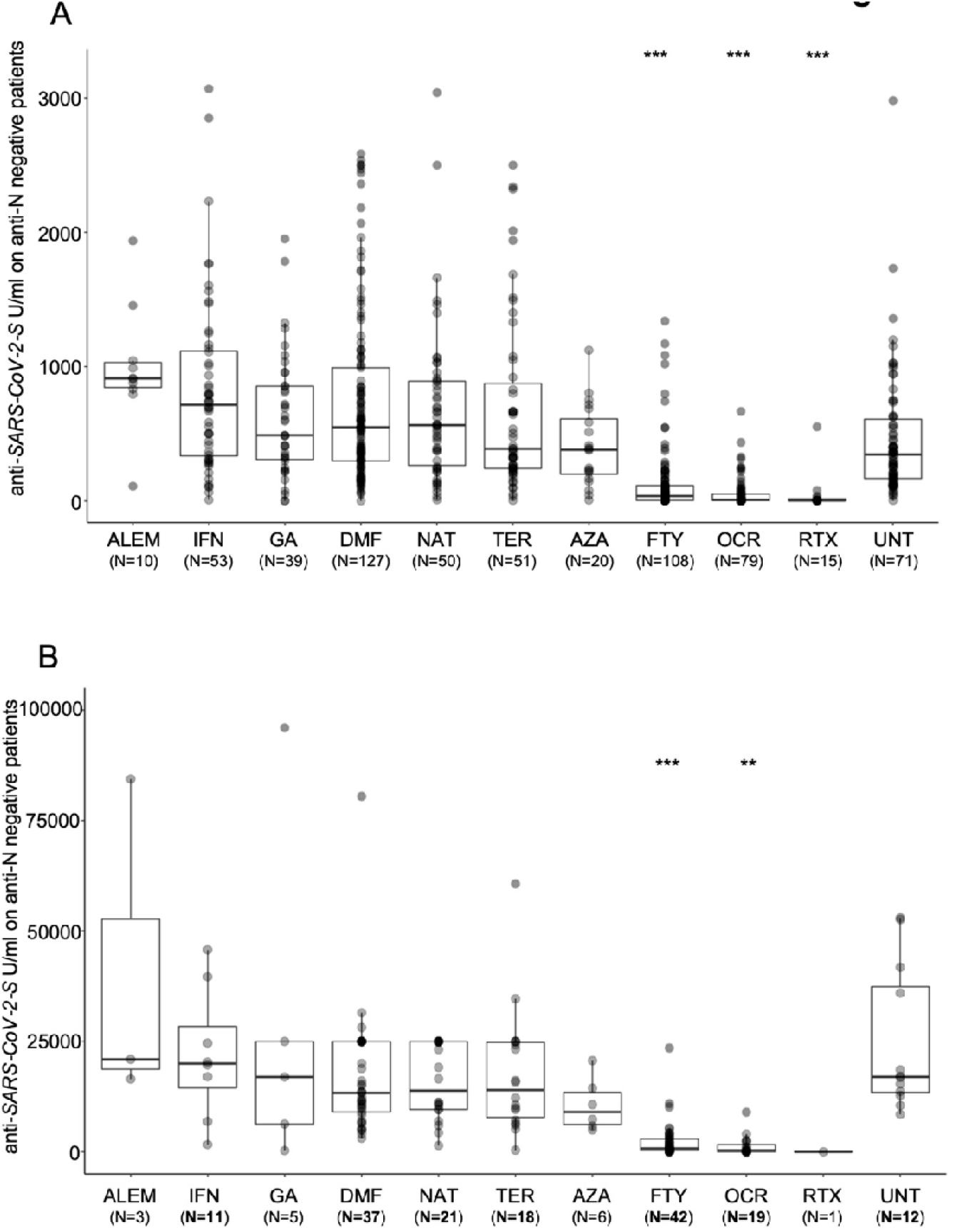
Post-vaccination SARS-CoV-2 anti-S antibody response by disease-modifying therapy (DMT) in MS patients negative for anti-N antibodies. (A) Antibody response to SARS-CoV-2 vaccination by DMT in MS patients six month after SARS-CoV-2 vaccination (T2). (B) Antibody response one month after booster (T3) by DMT in MS patients. Results are reported as boxplots, showing the median value (in bold) and the quartiles as box limits; whiskers at the top and bottom sides represent the overall maximum value and the overall minimum value, respectively. Data outside boxes and whiskers represent the outliers of the distribution. (*P< 0.05; **P<0.01; ***<0.001). (UNT=untreated; ALEM=alemtuzumab; IFN=interferon; GA=glatiramer acetate; DMF=dimethyl fumarate; NAT=natalizumab; CLA=cladribine; TER=terifluomide; AZA= azathioprine; FTY=fingolimod; RTX=rituximab; OCR=ocrelizumab)

The nucleocapsid protein antigens analysis, used to discriminate the immune response generated by the vaccination from immune response generated by natural SARS-CoV-2 infection, identified 23 anti-N positive patients at T2 (3.5% of 662 patients) and 9 at T3 (4.9% of 185 patients) (**Table 1**). Prior natural SARS-CoV-2 infection impact the humoral responses to BNT162b2 vaccine (2). Indeed, in MS patients with evidence of a natural exposure to SARS-CoV-2 virus, postvaccination anti-S antibodies levels were significantly higher than in patients who did not experience SARS-CoV-2 infection at T2 (medians 2,500 vs. 303.4 U/ml, Mann-Whitney test p = 2.36×10^−11^). We observed a comparable trend at T3 (medians 10,965 vs. 9,600 U/ml, Mann-Whitney test p = 0.4816). However, the number of MS patients positive for anti-N proteins was small, especially at T3, and additional data are required to properly evaluate the impact of SARS-CoV-2 infection on the immune response to BNT162b2 vaccine.

### The influence of disability status, sex, age, and smoking on anti-S production after BNT162b2 vaccine

We also tested at T2 and T3 the impact of additional factors that had been shown to influence anti-S production after BNT162b2 vaccine at T1 (2). In contrast with findings at T1, at T2 our statistical model did not reveal additional significant effects for sex, while age and the Expanded Disability Status Scale (EDSS) continue to show consistent effects. Specifically, we observed reduced postvaccination levels of anti-SARS-CoV-2 antibodies directed against the S protein in older patients (IRR = 0.98, p = 4.459×10^−07^) as well as in patients with reduced EDSS (IRR = 0.95, p = 0.022). At T3 only the reduced EDSS showed a significant effect (IRR = 0.89, p = 0.01) on anti-SARS-CoV-2 antibodies production.

Due to the impact of smoking on antibodies production in healthy and unhealthy cohorts of smokers compared to non-smokers (2,9), we examined the effects of active cigarette smoking on humoral response to SARS-CoV-2 vaccine in a subset of MS patients negative for anti-N antibodies production for whom smoking status was available (T2=510, T3=157). Our analyses showed that active cigarettes smoking reduced anti-S antibodies production (median = 128.0 U/ml) compared to non-smokers (median = 349.6 U/ml) in response to BNT162b2 vaccine (Mann-Whitney test p = 4.9×10^−4^) at T2. No differences were observed at T3 (Mann-Whitney test p = 0.3). The discrepancy between the three time points analyzed could be due to the effect of vaccination over time or the sample size, significantly smaller at T3 than T2.

### *Cross sectional and longitudinal study on anti-S and anti-N* SARS-CoV-2 *antibody levels*

Overall, evaluating the median of anti-S antibody level at each timepoint (T1-T2-T3) (**Figure 1**), we found that 6 months after the second dose of BNT162b2 vaccination (T2), the levels of anti-S decrease 3-fold compared to T1 (median T1= 962.2 U/ml and T2 = 323.7 U/ml). Furthermore, in line with previous reports (10), a strong upregulation in serum anti-S antibody levels was observed a month after the BNT162b2 booster (median T3 = 9,758 U/ml). The anti-S antibody median at T3 is ∼30-fold higher than T2 and ∼10-fold higher than T1. After stratification for each DMTs analyzed in this study, we observed that antibody levels significantly decrease when comparing T1 with T2 for the following treatments: interferon (IFN), DMF, natalizumab (NAT) and teriflunomide (TER) (**Figure 3**). Furthermore, the booster significantly increased anti-S production for DMF, NAT, TER, FTY and OCR. Interestingly, the increase in anti-S antibodies due to the booster was also observed for FTY and OCR even if with a lower impact (**Figure 3**). The upregulation of SARS-CoV-2 anti-S antibodies at 4 weeks after the booster (T3) was confirmed in an additional longitudinal analysis of a subgroup of 94 MS patients with data at every time points (Median T1= 888 U/ml, T2= 562 U/ml and T3 = 13,346 U/ml) (**Table 4**).

**Table 4.** Median of anti-S SARS-CoV-2 antibody levels in 94 MS patients enrolled at every time point (T1, T2 and T3) and stratified by DMTs.

| DMTs | N | Median anti-S in U/ml (IQR 25-75) |  |  |
| --- | --- | --- | --- | --- |
|  |  | T1 | T2 | T3 |
| ALEM | 3 | 1696 (1204-2297) | 833 (472-871) | 20942 (18717-52661) |
| AZA | 2 | 559 (368-750) | 565 (490-641) | 10894 (9132-12656) |
| DMF | 23 | 1859 (1065-3200) | 530 (293-766) | 13346 (8705-22508) |
| FTY | 21 | 45 (8-113) | 18 (4-156) | 555 (144-3937) |
| GA | 3 | 888 (466-2221) | 214 (132-685) | 16962 (8623-56476) |
| IFN | 5 | 1341 (862-1502) | 934 (500-2849) | 20335 (19660-132280) |
| NAT | 10 | 1700 (853-2787) | 619 (503-746) | 13577 (9596-23535) |
| OCR | 12 | 10 (8-405) | 6 (0.4-99) | 272 (8-978) |
| RTX | 1 | 8 (8-8) | 0.4 (0.4-0.4) | 8 (8-8) |
| TER | 8 | 557 (389-664) | 559 (176-661) | 11060 (6156-17912) |
| UNT | 6 | 2230 (1420-2835) | 647 (342-895) | 15406 (13028-31248) |

Abbreviations: UNT=untreated; ALEM=alemtuzumab; IFN=interferon; GA=glatiramer acetate; DMF=dimethyl fumarate; NAT=natalizumab; CLA=cladribine; TER=teriflunomide; AZA=azathioprine; FTY=fingolimod; RTX=rituximab; OCR=ocrelizumab; IQR= interquartile range.

**Figure 3.**
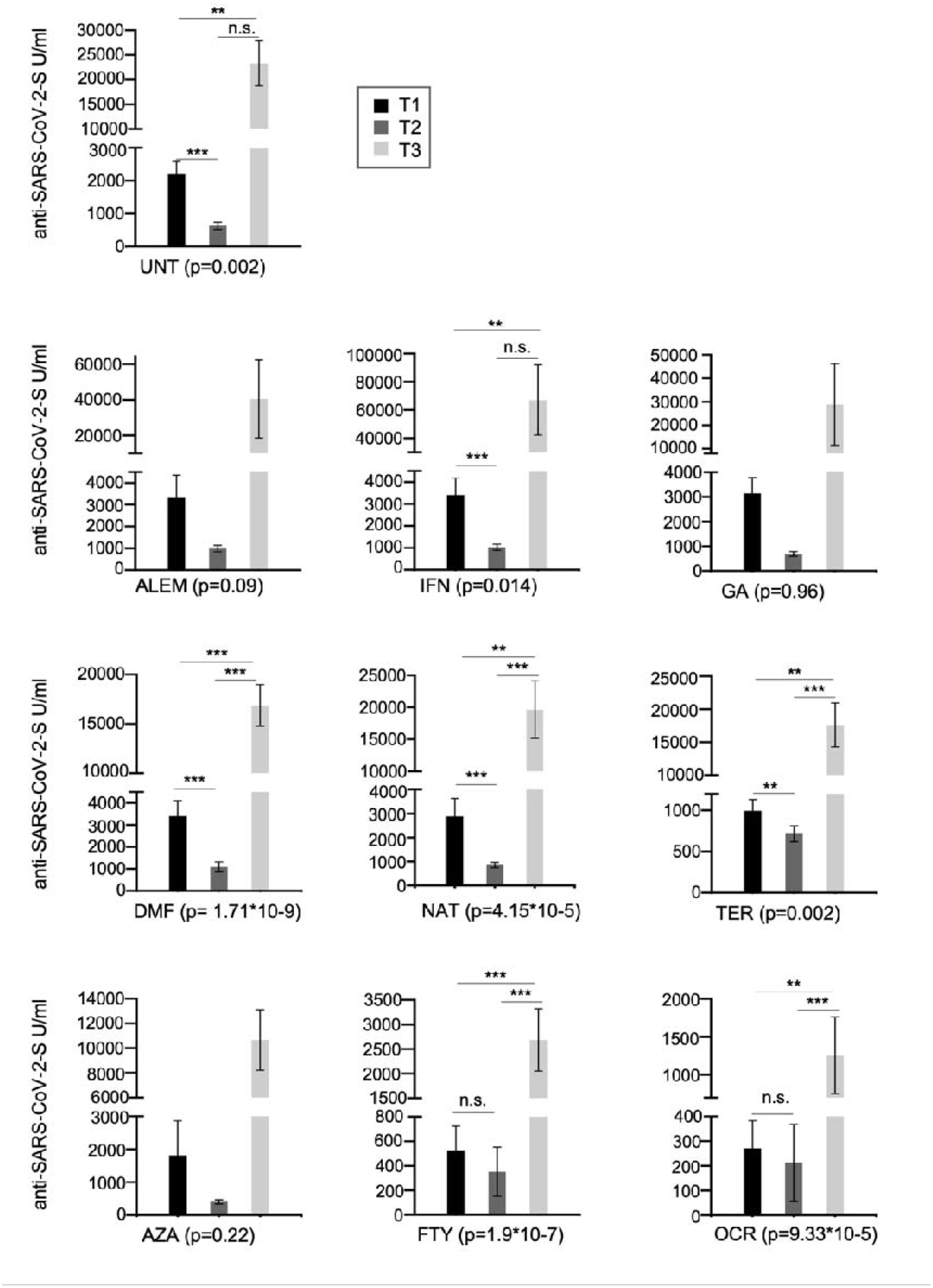
Cross-sectional humoral response over time (T1, T2 and T3) in MS patients treated with different DMTs or untreated. Post-vaccination anti-S antibody response by disease-modifying therapy (DMT) in MS patients. Results are represented as histogram showing the median value. Significance was tested using Mann-Whitney test. (*P< 0.05; **P<0.01; ***<0.001). (UNT=untreated; ALEM=alemtuzumab; IFN=interferon; GA=glatiramer acetate; DMF=dimethyl fumarate; NAT=natalizumab; CLA=cladribine; TER=terifluomide; AZA= azathioprine; FTY=fingolimod; RTX=rituximab; OCR=ocrelizumab)

Longitudinal analysis of anti-N antibodies also revealed a progressive decline in anti-N proteins from T1 to T3. Indeed, all 13 MS patients positive for SARS-CoV-2 infection at T1 (as determined by anti-N positivity), and with data for at least another time point, were anti-N negative ∼7-9 month later demonstrating a consistent loss of circulating humoral response against N antigens over time (data not shown). It is important to take this into account when testing samples without information on COVID-19 disease status. Indeed, if exposure occurred well before the time of antibody testing, it may be impossible to detect individuals with natural humoral immunity against the virus.

### Impact of different DMTs on COVID-19 risk and severity

To evaluate the risk of COVID-19 in patients receiving anti-CD20 (OCR and RTX) and FTY therapies, which are associated with reduced humoral responses to vaccine, we assessed the number of MS patients who became positive for COVID-19 five months after the booster. The presence of symptomatic infection and its severity was evaluated. A subset of 140 MS patients was monitored and among them 52 patients (37%) were treated with anti-CD20 or FTY while 88 (62.9%) received other therapies or were untreated. Overall, 28 MS patient (20%) of the 140 examined became positive for COVID-19 during the five months following the booster. Among them, COVID-19 infection was significantly more common (risk ratio = 1.95, 95% C.I =1.01 - 3.77, p=0.046) in patients under anti-CD20 and FTY therapies as compared to patients receiving other therapies or untreated. The higher incidence of disease in the group treated with anti-CD20 and FTY compared with the other therapies correlates with the lower antibody titers observed in those DMT categories. Importantly, regardless of the DMT received, all patients reported mild symptoms that did not require hospitalization.

## Discussion

Following previous observations on humoral responses to the BNT162b2 vaccine one month after the second dose of vaccine (T1) in MS patients treated with different DMTs or untreated (2), here we performed a cross-sectional and longitudinal follow-up study to analyze the humoral response in MS patients 6 months (T2) after the second dose and to 1 month after the first booster dose (T3) of BNT162b2vaccine.

In line with early results obtained at T1(2), we show that, compared to untreated patients, humoral responses in patients treated with FTY, OCR and RTX were impaired at T2 while in general they were not significantly different in patients treated with the other DMTs including alemtuzumab (ALEM), IFN, glatiramer acetate (GA), DMF, NAT, TER and AZA (**Table 2**).

Regarding the effects of the first booster dose of vaccine, at T3 we observed a strong upregulation of the humoral response in both treated and untreated patients, although the humoral responses observed in patients treated with anti-CD20 (OCR) and FTY remained significantly lower than those observed in untreated patients (**Table 3 and 4**). Yet, the first booster dose significantly increases anti-S levels even in most patients taking these immunosuppressive therapies. For example, in anti-N negative patients under OCR and FTY the median anti-S antibody levels were, respectively, 40.5 and 28.6-fold higher at T3 than at T1.

Likewise, prior natural SARS-CoV-2 infection, documented through the presence of anti-N antibodies, also strongly potentiated the humoral responses to BNT162b2 vaccine by inducing a strong increase in levels of anti-S antibodies at both T2 and T3 regardless of the DMTs analyzed and including, albeit to a lesser extent, patients treated with anti-CD20 or FTY.

Anti-N antibody analysis also provided additional data on the rate of decline in anti-N seropositivity which is essential for identifying patients with asymptomatic SARS-CoV-2 infection. We found that anti-N antibody levels declined at a sharper rate over time until they were no longer detectable, at least with the assay used in this study, 7 to 9 months after infection. Thus, it is possible that natural asymptomatic SARS-CoV-2 infections during the early phase of the pandemic were not detected in some of the MS patients considered here, and that these early infections still had a residual but not ascertainable effect on anti-S antibodies produced after vaccination. This is in agreement with a cross-sectional study by Alfego and colleagues that showed a decay rate of anti-N responses of 68.2% after only 9.7 months following infection, whereas anti-S seropositivity maintained a positivity rate of 87.8% 10 months later (11).

In addition to DMTs and prior SARS-CoV-2 infection, in agreement with previous result at T1 (2), we confirmed that both older age and reduced EDSS also influence antibody response to vaccine. Anti-S antibodies levels, however, were significantly lower in older patients at T2 but not a T3. The discrepancy between T1, T2 and T3 is probably due to the smaller sample size we analyzed at T3. Similar findings have been reported in a general population (12) where older age was associated with lower seroconversion rates. By contrast, Cohen and colleague have reported associations between older age and higher immune responses to natural infection, including IgG neutralizing antibody and memory B cell levels (13).

We also evaluated the relationship between antibodies titers in response to BNT162b2 vaccine and active smoking status in a subgroup of MS patients. Our data showed a significant downregulation in anti-S antibodies in MS patients who were active smokers, in line with the association of smoking with immune system dysfunctions (14).

An important preliminary observation of this study is the about two-fold higher risk of COVID-19 disease observed in the group treated with anti-CD20 and FTY compared with other therapies; a result which is somewhat expected given the lower antibody levels observed in those DMT categories. However, despite the higher incidence of COVID-19, these patients reported mild symptoms that did not require hospitalization, like those treated with other DMTs. This suggests that vaccination, and in particular booster doses of the vaccine, still may provide additional protection against severe COVID-19 disease most likely by enhancing immune responses like those mediated by T cells, that are not directly affected by these DMTs and by further stimulating residual B-cell and humoral responses. However, the sample size considered in this study, especially for patients treated with some DMTs, is small to draw firm conclusions. Our observations should be thus expanded to larger case series and further followed over time, including consideration of the immune and clinical impact of additional vaccine booster shots.

In this regard, the results presented here refer to the evaluation of the effects of the initial anti-Sars-Cov2 vaccination followed by a single booster dose of the vaccine completed several months ago and should therefore be placed in a more current context. Initial reports suggest that an additional booster - already approved by many regulatory agencies for the elderly and immunocompromised individuals - improves protection without affecting safety (15). Given these findings and the improved immune responses to vaccine after prior infection and/or the first booster dose of vaccine, it is likely that the use of further booster shots -- with existing mRNA vaccines and possible new generation of mRNA vaccines designed on emerging SARS-CoV-2 variants of concern -- will further improve immune responses against SARS-CoV-2 of MS patients under any DMT and thus their safety.

## Data Availability

All data produced in the present study are available upon reasonable request to the authors

## Study Funding

The study was supported by the Italian Foundation for Multiple Sclerosis-FISM (Grant N. 22021/Special/002 and 2021/C19-R-Single/010)

## Glossary

(MS): Multiple sclerosis
(DMTs): disease-modifying therapies
(SARS-CoV-2): severe acute respiratory syndrome coronavirus type-2
(COVID-19): Coronavirus disease 2019
(Ab): Antibody
(S): Spike
(N): Nucleocapsid

## Acknowledgment

We thank all the patients and volunteers who generously participated in this study. We would also like to thank all the medical doctors, nurses, and students who have collaborated to the realization of this study.

## Notes

### Competing Interest Statement

The authors have declared no competing interest.

### Funding Statement

This study was funded by FISM

### Author Declarations

Ethics committee of TS Sardegna gave ethical approval for this work

